# National Health Examination Surveys: an essential piece of the health planning puzzle

**DOI:** 10.1101/2023.07.11.23292221

**Authors:** Paula Margozzini, Hanna Tolonen, Antonio Bernabe-Ortiz, Sarah Cuschieri, Chiara Donfrancesco, Luigi Palmieri, Luz Maria Sanchez Romero, Jennifer S Mindell, Oyinlola Oyebode

**Affiliations:** Department of Public Health, Faculty of Medicine, Pontificia Universidad Católica de Chile, Santiago, Chile; Finnish Institute for Health and Welfare (THL), Helsinki, Finland; Universidad Científica del Sur, Lima, Peru; University of Malta, Msida, Malta; Istituto Superiore di Sanità, Rome, Italy; Georgetown University, Washington DC, USA; University College London, London, UK; Queen Mary University of London, London, UK

**Author notes:** Joint first (Paula Margozzini, Hanna Tolonen). Joint last (Jennifer S Mindell, Oyinlola Oyebode). **CRediT statement** Conceptualization JM Methodology CD, JM, PM, OO, LP, HT, Investigation SC, CD, ABO, PM, JM, LP, LMSR, HT, Writing - Original Draft JM, PM, OO, HT Writing - Review & Editing SC, CD, JM, PM, ABO, OO, LP, LMSR, HT, Supervision JM Project administration JM.

**Keywords:** Health examination survey, Objective health measurements, Policy, Evidence, Europe, Latin America

## Abstract

National health examination surveys (HESs) have been developed to provide important information that cannot be obtained from other sources. A HES combines information obtained by asking participants questions with biophysical measurements taken by trained field staff. They are observational studies with the highest external validity and make specific contributions to both population (public health) and individual health. Few countries have a track record of a regular wide-ranging HES, but these are the basis of many reports and scientific papers. Despite this, little evidence about HES usefulness and impact or the factors that influence HES effectiveness have been disseminated. This paper presents examples of HES contributions to society in both Europe and the Americas. We sought information by emailing a wide list of people involved in running or using national HESs across Europe and the Americas. We asked for examples of where examination data from their HES had been used in national or regional policymaking. We found multiple examples of HES data being used for agenda-setting, including by highlighting nutritional needs and identifying underdiagnosis and poor management of certain conditions. We also found many ways in which HES have been used to monitor the impact of policies and define population norms. HES data have also been used in policy formation and implementation. HES data are influential and powerful. There is need for global support, financing and networking to transfer capacities and innovation in both fieldwork and laboratory technology.

## Introduction

National health examination surveys (HESs) have been developed to provide important information that cannot be obtained from other sources. A HES combines information obtained by asking participants questions (whether through face-to-face interview, self-completion questionnaires, other modes, or a combination of these) with biophysical measurements taken by trained field staff. They are observational studies with the highest external validity and make specific contributions to both population (public health) and individual health.(1)

Unlike a health interview survey (HIS), where only self-reported information is obtained, a HES collects objective, biophysical measurements to complement self-reported data. This can be more accurate. For example, people tend to overestimate their height and underestimate their weight when asked, compared with measurements taken by trained staff, leading to underestimation of body mass index (BMI).(2–5)

Routine data from health services can sometimes be used to derive the evidence needed for policy-making. However, these data do not capture those who are not using health services. This is particularly problematic when information is needed on conditions that are often diagnosed late (or undiagnosed), such as diabetes. Unlike interviews and disease registers, a HES can quantify the prevalence of undiagnosed disease. For example: diabetes can be examined through measurement of glycated haemoglobin or fasting glucose; probable hypertension can be examined through blood pressure measurements.

Most high- and middle-income countries conduct national HIS to collect basic information about the general population using a questionnaire. For members of the European Union, it is mandatory to conduct the European Health Interview Survey: https://ec.europa.eu/eurostat/web/microdata/european-health-interview-survey. However far fewer countries conduct a regular HES. Many low- and middle-income countries have conducted at least one small HES following the World Health Organization (WHO)’s STEPwise Approach to Noncommunicable disease (NCD) Risk Factor Surveillance (STEPS) guidance: https://www.who.int/teams/noncommunicable-diseases/surveillance/systems-tools/steps. In addition, the programme of Demographic and Health Surveys (DHSs) funded by USAID are run at least 5-yearly in many LMICs and annually in others, e.g. Peru. These include a small number of biophysical measurements such as anthropometry and anaemia testing: https://dhsprogram.com/. Few countries have a track record of many HESs that include a wide range of bio-measures. The longest running HES in Europe is conducted in Finland, every five years since 1972 (FINRISK) (6); the USA’s National Health And Nutrition Examination Survey (NHANES) started in the 1960s, being continuous since 1999.

Existing national HESs vary by age-group covered; range of measurements taken; and organisation. For example most HESs in mainland Europe and in the USA use a clinical examination centre, while those in England and Latin America visit participants in their own homes.(7,8) However, they share many features despite some specific differences.

Despite the numerous reports and scientific papers reporting findings based on HES data, implicitly suggesting that these are valuable data sources, many countries still do not have a regular, nationally-representative, wide-ranging HES. In addition, those who have them, are struggling to secure long term financing for successive waves. Countries with small populations face additional challenges, including lack of infrastructure and resources to conduct a regular nationally-representative HES.(9)

Apart from a couple of papers from England (10,11) and Finland,(12) little evidence about HES usefulness and impact, or the factors that influence HES effectiveness has been disseminated; this needs to be communicated better to stakeholders. This paper will discuss the arguments that support their potential contributions to society, providing concrete evidence of their use in both Europe and the Americas.

## Methods

We sought information by emailing a wide list of people involved in running national HESs across Europe and the Americas. We asked for examples of where examination data from their HES had been used in national or regional policymaking. We also sought counterfactual examples highlighting where policies might have been developed, implemented or targeted differently, had HES data been considered. Within certain countries, authors also contacted colleagues working in national ministries or governmental agencies to ask for examples. We characterised each example by where it contributed within the policy-making cycle: agenda-setting (in which a problem or challenge that impacts the public is identified); policy formation (development of policy options and decisions taken); policy implementation (how the chosen policy option is put into effect); and policy evaluation (monitoring policy impacts to determine if a policy is achieving the intended goal, and if there are unintended consequences).

## Results

### Agenda setting

Health examination data has been used to identify or quantify population health issues that need addressing, providing justification for attention and policy decisions. Examples relating to nutrition include identification of persistent high levels of maternal anaemia and stunting in Peru, identified in the DHS, which informed the Children Malnutrition initiative in 2006, with a National Strategy for Poverty Reduction and Economic Opportunities (CRECER) in 2007.(13) The DHS data also helped to demonstrate the need to target social and nutritional programmes on rural areas.(14) Secondly, Finnish HES data from 2002 showed that population vitamin D levels were low. These data supported fortification of milk and fat spreads with vitamin D, which began in 2003.(12) Thirdly, neural tube defects (NTDs, e.g. spina bifida) are important congenital defects that can have serious impacts on health and independence. Adequate folic acid intake (400μg/d) can prevent 20-50% of cases. NHANES data showed that median levels of red blood cell folate among women aged 15-45y in 1988-94 in the USA were 160ng/mL, substantially below the recommended level (220ng/mL), with 38% having levels below this. In response, mandatory fortification of cereal grain products was implemented in January 1998; by 1999-2000, median levels were 264ng/mL.(15) Finally, in Italy, the assessment of 24 hour urine collection within the HES identified a higher daily salt consumption in the adult population than that recommended by the WHO.(16) As a result, the Ministry of Health in collaboration with non-governmental organizations undertook health promotion actions aimed at increasing awareness of the risks associated with excessive salt intake and to reformulate a wide range of products with reduced salt content through voluntary agreements with the food industry and artisan bakeries.

Health examination data can be used to predict future disease burden, for example objective anthropometry can be used to predict likely numbers of cases of diabetes, for example.(17) Comparison of body mass index (BMI) from measurements with responses to questions about screening for diabetes can also highlight weaknesses in current diabetes screening programmes.(18)

HES can identify under-diagnosis and can also monitor disease management, for example hypertension (19–21) and diabetes.(22) In Italy, in HES 2008-2012, direct measurement of blood pressure, cholesterol and glycaemia identified that 39% of men and 34% of adult women were unaware of their raised blood pressure; 38% and 42%, respectively, were unaware of probable hypercholesterolemia; and 39% and 29%, respectively, were unaware of probable diabetes.(23) The 2018-2019 HES provided evidence of improvements in awareness of raised blood pressure and in controlled hypertension.(24) In England in 2009/2010, 3% of adults aged 75+ reported having CKD but 32% of adults had stage 3-5 CKD according to the HES measurements.(25) In Mexico in 2016, the HES noted that 40% of adults with hypertension were undiagnosed.(26) In Malta the HIS 2015 reported the adult prevalence for diabetes at 8.5%, however the HES 2014-2015 reported the prevalence to be 10.3% with 4% being newly diagnosed.(27)

Finally, HES data may generate local hypotheses of association between population exposure to risk factors and chronic diseases, e.g., the need to study the local causes of the high levels of thyroid stimulating hormone (TSH) that have been found in Chile.(28)

### Policy formation

The 2020 obesity strategy in England looked to ban TV and online advertising of food high in fat, sugar and salt (HFSS) before 21:00; restrict price promotions for HFSS foods; restrict where shops may display HFSS foods and drinks and mandate energy labelling of food in restaurants. These were assessed during development of the obesity strategy by using Health Survey for England anthropometric data in a Department of Health and Social Care ‘Calorie Model’ that explored likely impacts of those and others possible policy interventions https://assets.publishing.service.gov.uk/government/uploads/system/uploads/attachment_data/file/736417/dhsc-calorie-model-technical-document.pdf. In Malta, anthropometric data used to accurately estimate obesity have also been useful, these data contributed to the cost estimation of this disease at a national level. This exercise was done to engage different stakeholders with the aim to activate joint working, in a timely fashion for the prevention of obesity.(29)

HES data has fed into guideline development. For example in the USA, the 2018 Physical Activity guidelines differed from the 2008 guidance in dropping the requirement for aerobic activity to be performed in episodes of at least 10 minutes https://health.gov/sites/default/files/2019-09/Physical_Activity_Guidelines_2nd_edition.pdf. This was informed by the 2003-2006 NHANES accelerometer data, linked to deaths data, showing that bout length was not important for mortality benefits of moderate-to-vigorous physical activity accumulated.(30)

HES data has supported decision making including where policies should be targeted. The Department for Health in England worked with the food industry to reduce salt content in processed food products that contributed most to the population’s salt intake, identified through health examination data.(31) For this reason, initial reformulation focused on bread. HES data highlighted the stark difference in prevalence of allergies between rural and urban populations in Finland, suggesting a loss of protective factors for urban residents. This prompted a policy approach within the Finnish Allergy Programme 2008-2018, based on increasing connection of the population to natural environments which has been successful.(32)

Modelling based on Chilean HES data was used by the Ministry of Health to persuade the Treasury to expand the range of treatments covered for endstage CKD (M. Walbaum, personal communication).

### Policy implementation

FINRISK HES data linked to national healthcare datasets were used to create the FINRISK calculator to identify individuals at high risk of cardiovascular disease (33) and the CAIDE score to predict subsequent development of dementia.(34) The data were also used to validate the FINDRISC tool for identifying high diabetes risk.(35) Chilean HES data linked to national hospital discharges and deaths were used to assess the predictive validity of a new multimorbidity stratification scheme that is now being scaled to primary care at the national level (Margozzini P, personal communication).

### Policy monitoring and evaluation

Repeated health examination measures can be used to monitor changes in health status over time, and this means it is possible to see whether implemented policies are having the desired effects. Following on from the examples of agenda-setting reported above, in Peru, the policies to address child malnutrition, alongside a cash transfer programme are credited with the improvements seen in nutritional status.(36) In Mexico, ENSANUT data demonstrated that the Social Food Assistance Programs reduced food insecurity (37). In Finland, the national HESs were used to examine whether vitamin D fortification supported better population blood levels. Mean serum 25-hydroxyvitamin D levels rose from 48nmol/l in 2000 to 65nmol/l in 2011 (p<0.001), with a higher rise of 20nmol/l in survey participants not taking supplements.(38) By 1999-2000, NHANES data showed a dramatic increase in folate levels to a median of 265ng/mL. Birth rates of babies born with NTDs fell in the two years prior to mandatory fortification, probably due to increased rates of folate supplementation and prenatal diagnosis, and fell further in the two years after mandatory fortification. The NTD rates continued to fall, more slowly, although median folate levels fell after 2000.(15) In Italy, to evaluate the effectiveness of the population salt reduction strategy implemented by the Italian Ministry of Health in collaboration with non-governmental organizations, monitoring of the adult population salt intake was done through the collection of 24hour urine samples. This showed a reduction of about 12% of sodium urine excretion from 2008/12 to 2018/19.(16) Similar monitoring was implemented regarding the consumption of potassium, an important indicator of fruit and vegetable consumption.(39)

Other similar examples include the DHS being used to monitor the double burden of malnutrition and policies to address this in Mexico (40). 24 hour urine collection was also done to monitor salt reduction policies in England, but despite targets set in 2006, no significant falls in salt intake were seen from 2008/09 to 2018/19.(41) Chilean Encuesta Nacional de Salud (ENS i.e.: National Survey of Health) has helped to monitor a change in the mandatory fortification programmes in reaction to the ongoing nutritional transition.(42)

In the US, NHANES measurements of blood lead levels have shown falls over time in adults and children, including in high-risk areas, attributed to Federal policies and a range of public health interventions to reduce exposure to lead (https://www.cdc.gov/nceh/lead/data/nhanes.htm).

Even where measurements aren’t used directly, they can be used to improve survey data. Measurement data for height and weight have been used by Public Health England to correct self-reported anthropometric data from the Active People’s Survey in England to create the adult excess weight indicator in the Public Health Outcomes Framework.(43) The Active People Survey has 1,000 participants from each local government area; that level of coverage would be far too expensive for a HES, but the HES support interpretation of the self-reported interview data.

### Population norms

HES data have been used for defining population norms or reference values for the population regarding multiple diagnostic and laboratory tests (e.g., blood tests to detect hypothyroidism, liver enzymes, vitamin levels, urinary iodine excretion, cut-off points for waist circumference).(44) National Health and Nutrition Examination Survey (NHANES) measurements in the USA were the basis for the 2000 Center for Disease Control and Prevention Growth Charts for children that updated the 1977 National Center for Health Statistics charts and created new body mass index-for-age (BMI-for-age) charts.(45) Such population level data can then be used to identify outliers. In the USA, blood levels of lead have been measured in the population as part of NHANES. The ‘blood lead reference value’ identifies children with blood lead levels above the 97.5^th^ percentile for children aged 1-5y (https://www.cdc.gov/nceh/lead/data/nhanes.htm).

## Discussion

In this paper, we have provided examples of the range of ways HES data have been used in policy-making, showing that these data are particularly useful for identifying issues amenable to policy intervention and for monitoring or evaluating national policy outcomes. It is considerably more expensive to run a HES than an interview survey, but it supports “best buy health policy” which will be more cost effective. One ‘cost-of-illness’ appraisal in England found that even if the most expensive HES options were considered, the cost of a HES monitoring chronic obstructive pulmonary disease (COPD), cardiovascular disease (CVD), hypertension, diabetes and obesity was less than 0.01% of the annual societal costs for those diseases (S Morris, personal communication). Yet, with strategic planning a HES can be conducted with minimal resources, as noted in Malta.(46)

A Finnish policy-maker said: *“Without the evidence-based information health policy would be just “guessing policy.”*.(12) This is exemplified by the first public health policy for England, *Health of the Nation*, published in 1992, which set a target of reducing obesity in adults aged 16-64y to 6% in men and 8% in women by 2005, in the absence of any trend data from examination surveys. In fact, 22% of men and 23% of women measured in HSE 2005 had obesity. Paucity of existing data meant the targets had been set without appreciating the rate at which obesity was increasing.(10)

In order to reduce the preventable and avoidable burden of morbidity, mortality and disability due to NCDs, the World Health Organization (WHO) Global Action Plan for the prevention and control of NCDs 2013-2020 was approved. for which in 2021, WHO launched an interim assessment of implementation and disseminated an implementation roadmap 2023-2030. The action plan provides for the Member States to achieve nine voluntary objectives that should be monitored using 25 population health indicators, many of which require direct measurements (e.g., obesity, blood pressure, salt excretion).

HES fill a need. A big part of the natural history of disease is hidden inside households and is absent from health records. Health records usually do not have subjective information that HES may collect as part of their questions (e.g. health perception, beliefs, attitudes, quality of life, lifestyles). Healthcare data generally record limited demographic and socio-economic details and little information on health-related behaviours – and those data may not be extractable from routine data without hand-searching of individual records. Clerical staff who complete health records are not always well trained; are dependent on the notes made by clinical staff, which may be incomplete; and lack a standardization process, so they often introduce bias or important observer variability. Most importantly, health records only cover those seeking medical care, and therefore include neither asymptomatic nor undiagnosed cases which can be identified in a HES.

Other, non-traditional, electronic datasets are also hailed as potential sources of population health data.(47) However, these share the problems of healthcare data of excluding asymptomatic individuals alongside the biases due to the non-random nature of those providing information. In particular, those from poorer households and older people have less access to the internet or electronic devices, so are more likely to be missed by non-traditional data.

Where there is a lack of national HES data, policy makers are compelled to resort to other types of data, such as health records, self-reported HIS data, registries, and small population studies carried out by different researchers when formulating national policies. Such data fragmentation typically prevents the ability of data linkage, resulting in an incomplete and potentially inaccurate data assessment through overinflation or underreporting of the health problem which can mis-represent true epidemiology of key health issues. For example, while there is an established gradient in which non-communicable diseases are most prevalent among the most socio-economically deprived in high-income countries; in low and middle-income countries relying on self-reported diagnoses to examine non-communicable disease morbidity may lead to an artefactual association in which morbidity appears greatest in wealthier populations. Hence, relying on these types of data poses the risk of formulating strategies that do not target the realistic national health burden.(48) Additionally, the lack of national HES data presents as a challenge to quantify the impact of a disease on the population through national burden of disease studies. Without these studies, the true burden of a disease through morbidity and mortality cannot be measured; this will distort the true national problem, resulting in policymakers not having adequate data to act appropriately. Using HES data linked to healthcare, disease register, or mortality data can yield estimates of the proportion of disease, death, or healthcare use attributable to particular health-related behaviours identified using objective measures, such as cotinine to assess active and/or passive smoking or accelerometery to assess physical activity. This can also be used to conduct national burden of disease studies even for small population countries.(49)

By involving many international experts on HESs, including from two international networks in writing this paper, we hope to have captured the key ways in which HES data are useful in policy-making. However, this is not a systematic study of all the instances of use of HES data. In fact, conversations with policy-makers in the then Department of Health in England during a previous, systematic study (10,11) revealed that many examples of policy use of HES data are not documented, thus a comprehensive review is not possible. In addition, we have not sought to capture all the ways that HES data are used beyond policy-making, for example, in research and for development and training of undergraduate and graduate researchers. This article captures examples of the usefulness of HES for direct policy-making, as funding usually comes from government.

## Conclusions

HESs are influential sources of data and particularly powerful for identifying health issues and monitoring the impact of health policy. HES impacts should be better measured and communicated. There is need for global support, financing and networking to transfer capacities and innovation in both fieldwork and lab technology.

## Data Availability

There is no dataset for this study. Further details of any specific examples cited can be provided by the authors to interested readers.

## Funding and acknowledgements

No funding was received to undertake this study or prepare this manuscript. LMSR is supported by USA National Cancer Institute of the National Institutes of health under Award Number K01CA260378. Neither that funder nor the funders of the health examination surveys played any role in decisions about writing or publishing this paper nor in its drafting or approval.

We should like to thank all those who provided examples to the research team. We also thank Dr. J. Jaime Miranda for his support in connecting the authors from Peru with the larger collaborative network. Finally, we thank the participants of all the health examination surveys mentioned, the field staff who carried them out, and the commissioners who funded them.

